# Change in Finances, Peer Access, and Mental Health Among Trans and Non-binary People in Canada During COVID-19

**DOI:** 10.1101/2021.12.13.21267077

**Authors:** Monica A. Ghabrial, Ayden I. Scheim, Caiden Chih, Heather Santos, Noah James Adams, Greta R. Bauer

## Abstract

**Purpose:** COVID-related stressors associated with loss of income and community are compounded with gender minority stress among trans and non-binary people (TNB) – which may result in mental health burden. The present study examined the effect of COVID-related change in finances and TNB gathering access on anxiety and depression among TNB people.

**Methods:** Participants were 18 years and older (*M* age = 30) who completed both pre-pandemic baseline (Fall 2019) and pandemic follow-up (Fall 2020) surveys in the Trans PULSE Canada study. Multivariable regression analyses examined associations between change in (1) finances and (2) access to TNB peers and mental health during the pandemic.

**Results:** Of 780 participants, 50% reported that COVID had a negative effect on personal income and 58.3% reported loss in access to TNB peer or friend gatherings (in person or online). Depression and anxiety symptoms increased from pre-pandemic to follow-up, and most participants were above measurement cut-offs for clinical levels during the pandemic. Changes in finances and access to peer gatherings were associated with depression symptoms during the pandemic, but effects depended on level of pre-pandemic depression. For participants with high pre-pandemic depression, financial stability was not protective against increased depression at follow-up. Participants experiencing unprecedentedly high levels of depression during COVID may have pursued more TNB gatherings. Neither financial change nor access to TNB gatherings were associated with pandemic anxiety.

**Conclusion:** Findings suggest need for a multifaceted approach to mental health programmes and services to address structural barriers, including financial support and meaningful TNB community engagement.

## Introduction

The onset of the COVID-19 pandemic brought about abrupt closures, job loss, social distancing measures, and other interruptions to daily life [1]. These interruptions increased social isolation, housing and food insecurity, and stress at a population level [2,3]. These stressors have contributed to a deterioration in population mental health [3]. Trans and non-binary (TNB) people – those whose gender identity is different from their sex assigned at birth – already experience high levels of anxiety and depression, [4,5,6] which may have been exacerbated by the COVID-19 pandemic.

TNB individuals faced a disproportionate amount of economic and financial insecurity prior to the COVID-19 pandemic due to stigma and discrimination [7,8]. Many TNB people face employment discrimination and hostile work environments [9]. In the United States, 139,700 transgender adults were unemployed at the start of the pandemic and 96,400 experienced homelessness in the past year [7]. The extensive economic disruptions of the COVID-19 pandemic likely widened the financial gap between TNB and cisgender people. These structural vulnerabilities also increase the financial difficulties experienced by TNB with regards to affording gender-affirming surgeries, hormone medications, and supplies. This, in turn, can have a direct impact on mental health [5,10].

Social support, including that from family and peers, may serve to buffer the minority stressors encountered by TNB people [11]. Transgender adolescents, for example, have utilized social media platforms as community hubs to provide and receive emotional and informational support [12]. Another study reported that male-to-female transgender adults developed gender-based social networks through their community-based health care clinic, allowing them to build social capital and take political action [13]. Transgender individuals with low social support were also found to have the highest likelihood of discrimination associated suicidal ideation compared to those with moderate or high support [14]. However, the disruption of day-to-day activities caused by COVID-19 restricted access to community, social gatherings, friends, and other sources of support [10,15]. Compared to cisgender youth, TNB youth reported more mental health and substance use service disruptions along with less social support amidst the pandemic [15].

The objective of this article was to determine if self-reported changes in finances and TNB gathering access were associated with depression and anxiety symptoms among TNB individuals during COVID-19, using data from Trans PULSE Canada (TPC). We also assessed whether this relationship was moderated by pre-pandemic mental health symptoms.

## Methods

### Sample and Procedure

Trans PULSE Canada (TPC) was a community-based, national study of health and wellbeing among TNB people. Pre-pandemic data were collected over ten weeks in 2019 from 2,873 Canadian residents aged 14 and older, who were recruited using a multi-mode convenience sampling approach. A follow-up survey examining the impacts of COVID-19 pandemic was completed by 820 participants in Fall 2020. The 2019 survey could be completed in English or French online, by mail, by telephone, or in-person with a Peer Research Associate in 11 major urban areas. TPC and the COVID follow-up study were promoted on social media, email listservs, at community agencies and events targeted toward sexual and gender minority people, and through Peer Research Associate outreach. Participants were given the option of completing the full (∼60-minutes) or short-form (∼10 minutes) survey.

TPC was adapted from Trans PULSE – a community-based study on TNB health in Ontario, Canada in 2009-2010 – and was modified through an intensive consultation process by 9 priority population teams (trans people who are Indigenous, gender non-binary, racialized, sex workers, older adults, youth, northern and/or rurally living, disabled, and immigrants, refugees, and/or newcomers). In September and October 2020, participants who had consented to recontact (*n* = 1187) were invited to participate in the TPC COVID Cohort follow-up. This follow-up survey was programmed into REDCAP in English and French. Participants who completed the COVID follow-up received a $20 gift card. TPC was approved by the research ethics boards of Western University, Unity Health Toronto, Wilfrid Laurier University and the University of Victoria. The TPC COVID Cohort follow-up was approved at Western University, Wilfrid Laurier University, University of Victoria, Universal Health Network, University of British Columbia, and Drexel University.

This paper uses data collected prior to the pandemic and at the COVID-19 follow-up from participants who completed both stages of data collection and were over the age of 18 at baseline. We set the age minimum at 18 due to our focus on financial change during the pandemic, with the postulation that youth under 18 are more likely to be financially dependent on a guardian(s) and thus less likely to experience personal financial loss that would limit the ability to meet their needs. This resulted in 780 eligible participants.

### Measures

#### Sociodemographic variables

Participants responded to questions regarding social and demographic characteristics, including age, sex assigned at birth, immigration history, province, and details related to educational background, employment, and living situation. Racialization was assessed using two items: participants were asked if they identify as a person of Color and if they are perceived or treated as a person of Color in Canada. Individuals who responded with yes to either item were coded as racialized and those who did not were coded as non-racialized. Individuals who reported being First Nations, Inuit, Métis or otherwise Indigenous in Canada were coded as Indigenous, and all other participants were coded as non-Indigenous. Participants were asked to select a current gender identity from the following options: (1) man, (2) woman, (3) Indigenous or other cultural identity, (4) non-binary, genderqueer, agender, or similar.

#### Income and needs

Participants reported income using provided categories (see Table 1). We calculated estimated numerical income for each participant by assigning the mid-point of their selected income category. Income-to-needs ratio was calculated by dividing this numerical value by number of income dependents. To determine low-income category (Yes/No), we compared numerical income and number of dependents to the Statistics Canada low-income measure threshold for households, which is calculated using the number of dependents. For example, in 2019, the low-income measure threshold for a one-person household was $28,831 [16].

**Table 1.**
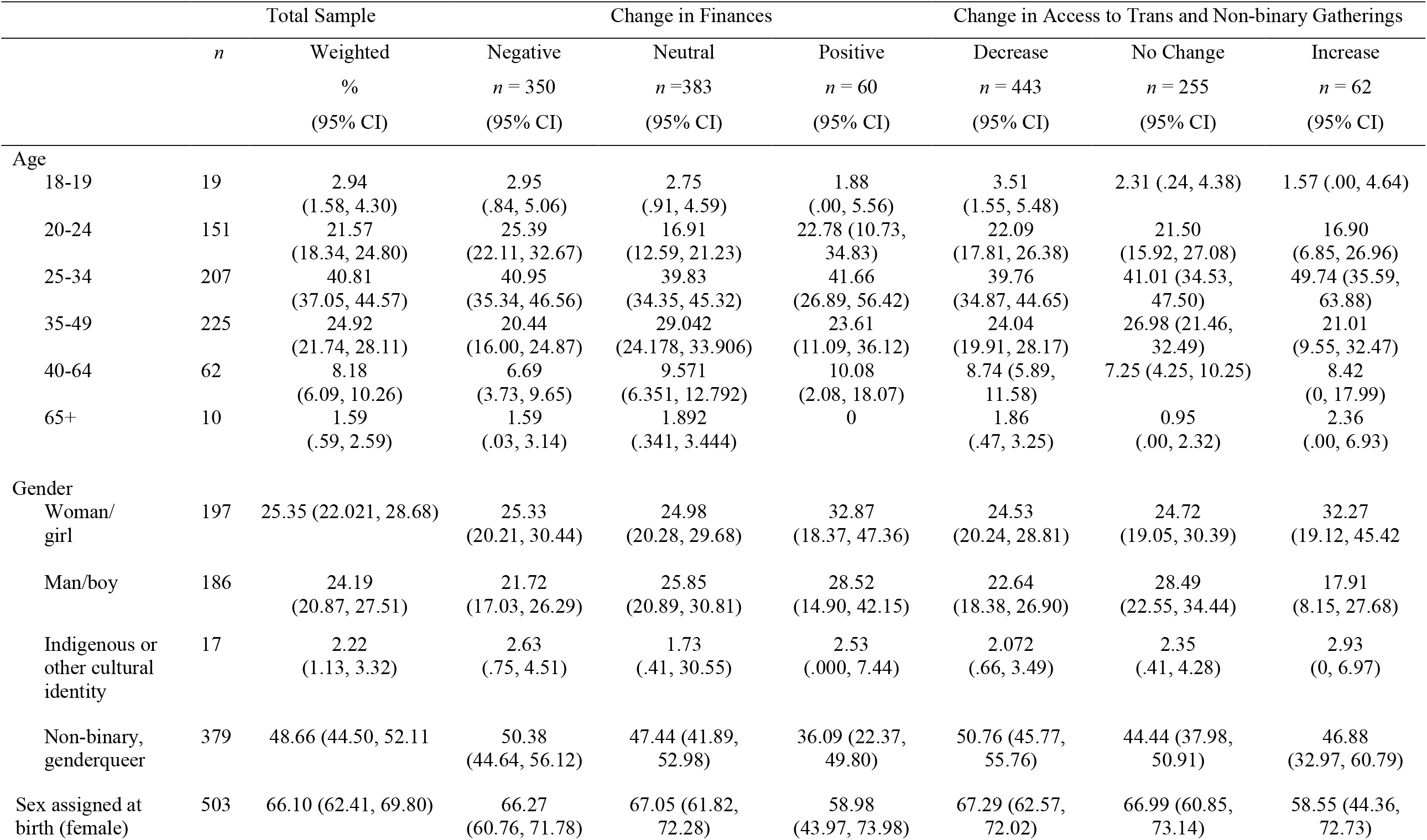
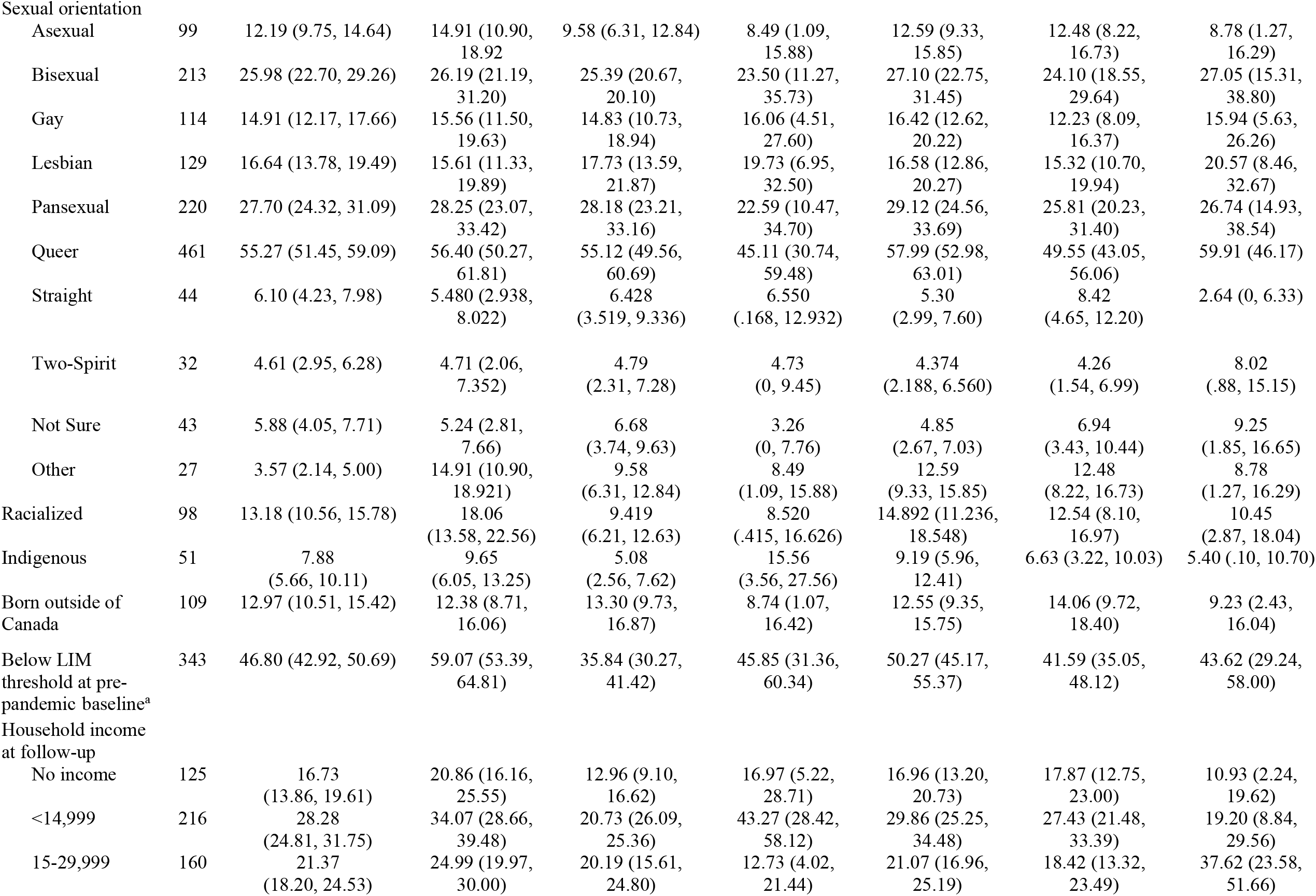
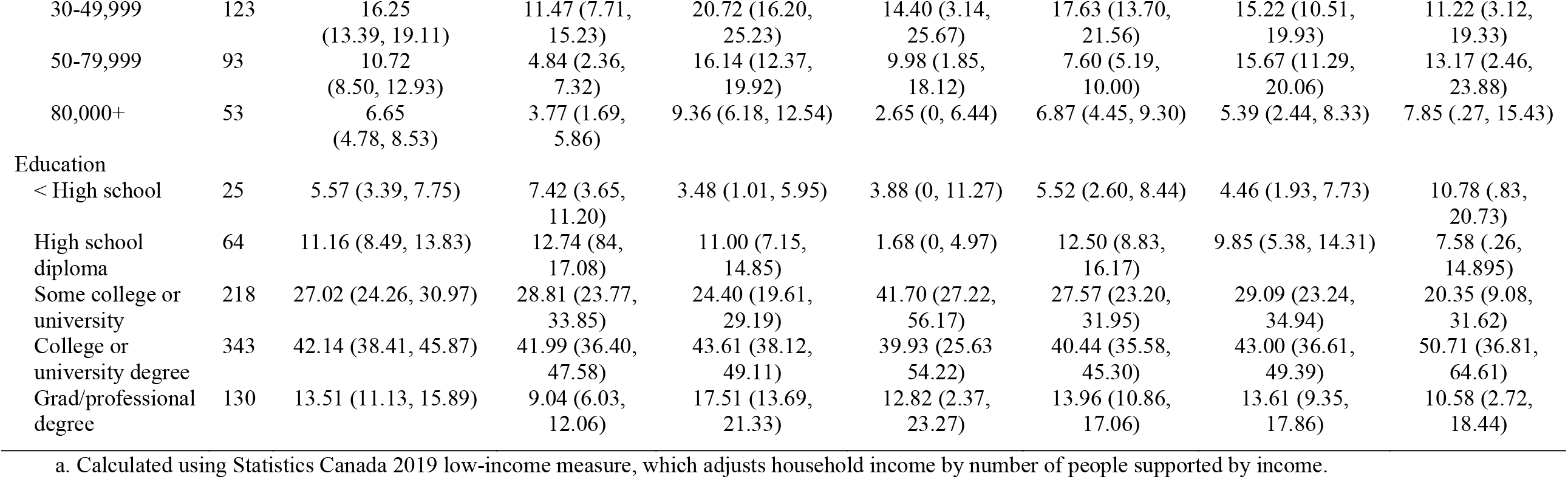
Participant characteristics, stratified by change in finances and access to trans and non-binary gatherings (*n* = 780)

|  | Total Sample |  | Change in Finances |  |  | Change in Access to Trans and Non-binary Gatherings |  |  |
| --- | --- | --- | --- | --- | --- | --- | --- | --- |
|  | *n* | Weighted<br>%<br>(95% CI) | Negative<br>*n* = 350<br>(95% CI) | Neutral<br>*n* = 383<br>(95% CI) | Positive<br>*n* = 60<br>(95% CI) | Decrease<br>*n* = 443<br>(95% CI) | No Change<br>*n* = 255<br>(95% CI) | Increase<br>*n* = 62<br>(95% CI) |
| Age |  |  |  |  |  |  |  |  |
| 18-19 | 19 | 2.94<br>(1.58, 4.30) | 2.95<br>(.84, 5.06) | 2.75<br>(.91, 4.59) | 1.88<br>(.00, 5.56) | 3.51<br>(1.55, 5.48) | 2.31 (.24, 4.38) | 1.57 (.00, 4.64) |
| 20-24 | 151 | 21.57<br>(18.34, 24.80) | 25.39<br>(22.11, 32.67) | 16.91<br>(12.59, 21.23) | 22.78 (10.73,<br>34.83) | 22.09<br>(17.81, 26.38) | 21.50<br>(15.92, 27.08) | 16.90<br>(6.85, 26.96) |
| 25-34 | 207 | 40.81<br>(37.05, 44.57) | 40.95<br>(35.34, 46.56) | 39.83<br>(34.35, 45.32) | 41.66<br>(26.89, 56.42) | 39.76<br>(34.87, 44.65) | 41.01 (34.53,<br>47.50) | 49.74 (35.59,<br>63.88) |
| 35-49 | 225 | 24.92<br>(21.74, 28.11) | 20.44<br>(16.00, 24.87) | 29.042<br>(24.178, 33.906) | 23.61<br>(11.09, 36.12) | 24.04<br>(19.91, 28.17) | 26.98 (21.46,<br>32.49) | 21.01<br>(9.55, 32.47) |
| 40-64 | 62 | 8.18<br>(6.09, 10.26) | 6.69<br>(3.73, 9.65) | 9.571<br>(6.351, 12.792) | 10.08<br>(2.08, 18.07) | 8.74 (5.89,<br>11.58) | 7.25 (4.25, 10.25) | 8.42<br>(0, 17.99) |
| 65+ | 10 | 1.59<br>(.59, 2.59) | 1.59<br>(.03, 3.14) | 1.892<br>(.341, 3.444) | 0 | 1.86<br>(.47, 3.25) | 0.95<br>(.00, 2.32) | 2.36<br>(.00, 6.93) |
| Gender |  |  |  |  |  |  |  |  |
| Woman/<br>girl | 197 | 25.35 (22.021, 28.68) | 25.33<br>(20.21, 30.44) | 24.98<br>(20.28, 29.68) | 32.87<br>(18.37, 47.36) | 24.53<br>(20.24, 28.81) | 24.72<br>(19.05, 30.39) | 32.27<br>(19.12, 45.42) |
| Man/boy | 186 | 24.19<br>(20.87, 27.51) | 21.72<br>(17.03, 26.29) | 25.85<br>(20.89, 30.81) | 28.52<br>(14.90, 42.15) | 22.64<br>(18.38, 26.90) | 28.49<br>(22.55, 34.44) | 17.91<br>(8.15, 27.68) |
| Indigenous or<br>other cultural<br>identity | 17 | 2.22<br>(1.13, 3.32) | 2.63<br>(.75, 4.51) | 1.73<br>(.41, 30.55) | 2.53<br>(.000, 7.44) | 2.072<br>(.66, 3.49) | 2.35<br>(.41, 4.28) | 2.93<br>(0, 6.97) |
| Non-binary,<br>genderqueer | 379 | 48.66 (44.50, 52.11) | 50.38<br>(44.64, 56.12) | 47.44 (41.89,<br>52.98) | 36.09 (22.37,<br>49.80) | 50.76 (45.77,<br>55.76) | 44.44 (37.98,<br>50.91) | 46.88<br>(32.97, 60.79) |
| Sex assigned at<br>birth (female) | 503 | 66.10 (62.41, 69.80) | 66.27<br>(60.76, 71.78) | 67.05 (61.82,<br>72.28) | 58.98<br>(43.97, 73.98) | 67.29 (62.57,<br>72.02) | 66.99 (60.85,<br>73.14) | 58.55 (44.36,<br>72.73) |
| Sexual orientation |  |  |  |  |  |  |  |  |
| Asexual | 99 | 12.19 (9.75, 14.64) | 14.91 (10.90, 18.92) | 9.58 (6.31, 12.84) | 8.49 (1.09, 15.88) | 12.59 (9.33, 15.85) | 12.48 (8.22, 16.73) | 8.78 (1.27, 16.29) |
| Bisexual | 213 | 25.98 (22.70, 29.26) | 26.19 (21.19, 31.20) | 25.39 (20.67, 20.10) | 23.50 (11.27, 35.73) | 27.10 (22.75, 31.45) | 24.10 (18.55, 29.64) | 27.05 (15.31, 38.80) |
| Gay | 114 | 14.91 (12.17, 17.66) | 15.56 (11.50, 19.63) | 14.83 (10.73, 18.94) | 16.06 (4.51, 27.60) | 16.42 (12.62, 20.22) | 12.23 (8.09, 16.37) | 15.94 (5.63, 26.26) |
| Lesbian | 129 | 16.64 (13.78, 19.49) | 15.61 (11.33, 19.89) | 17.73 (13.59, 21.87) | 19.73 (6.95, 32.50) | 16.58 (12.86, 20.27) | 15.32 (10.70, 19.94) | 20.57 (8.46, 32.67) |
| Pansexual | 220 | 27.70 (24.32, 31.09) | 28.25 (23.07, 33.42) | 28.18 (23.21, 33.16) | 22.59 (10.47, 34.70) | 29.12 (24.56, 33.69) | 25.81 (20.23, 31.40) | 26.74 (14.93, 38.54) |
| Queer | 461 | 55.27 (51.45, 59.09) | 56.40 (50.27, 61.81) | 55.12 (49.56, 60.69) | 45.11 (30.74, 59.48) | 57.99 (52.98, 63.01) | 49.55 (43.05, 56.06) | 59.91 (46.17, 73.66) |
| Straight | 44 | 6.10 (4.23, 7.98) | 5.480 (2.938, 8.022) | 6.428 (3.519, 9.336) | 6.550 (.168, 12.932) | 5.30 (2.99, 7.60) | 8.42 (4.65, 12.20) | 2.64 (0, 6.33) |
| Two-Spirit | 32 | 4.61 (2.95, 6.28) | 4.71 (2.06, 7.352) | 4.79 (2.31, 7.28) | 4.73 (0, 9.45) | 4.374 (2.188, 6.560) | 4.26 (1.54, 6.99) | 8.02 (.88, 15.15) |
| Not Sure | 43 | 5.88 (4.05, 7.71) | 5.24 (2.81, 7.66) | 6.68 (3.74, 9.63) | 3.26 (0, 7.76) | 4.85 (2.67, 7.03) | 6.94 (3.43, 10.44) | 9.25 (1.85, 16.65) |
| Other | 27 | 3.57 (2.14, 5.00) | 14.91 (10.90, 18.921) | 9.58 (6.31, 12.84) | 8.49 (1.09, 15.88) | 12.59 (9.33, 15.85) | 12.48 (8.22, 16.73) | 8.78 (1.27, 16.29) |
| Racialized | 98 | 13.18 (10.56, 15.78) | 18.06 (13.58, 22.56) | 9.419 (6.21, 12.63) | 8.520 (.415, 16.626) | 14.892 (11.236, 18.548) | 12.54 (8.10, 16.97) | 10.45 (2.87, 18.04) |
| Indigenous | 51 | 7.88 (5.66, 10.11) | 9.65 (6.05, 13.25) | 5.08 (2.56, 7.62) | 15.56 (3.56, 27.56) | 9.19 (5.96, 12.41) | 6.63 (3.22, 10.03) | 5.40 (.10, 10.70) |
| Born outside of Canada | 109 | 12.97 (10.51, 15.42) | 12.38 (8.71, 16.06) | 13.30 (9.73, 16.87) | 8.74 (1.07, 16.42) | 12.55 (9.35, 15.75) | 14.06 (9.72, 18.40) | 9.23 (2.43, 16.04) |
| Below LIM threshold at pre-pandemic baseline<sup>a</sup> | 343 | 46.80 (42.92, 50.69) | 59.07 (53.39, 64.81) | 35.84 (30.27, 41.42) | 45.85 (31.36, 60.34) | 50.27 (45.17, 55.37) | 41.59 (35.05, 48.12) | 43.62 (29.24, 58.00) |
| Household income at follow-up |  |  |  |  |  |  |  |  |
| No income | 125 | 16.73 (13.86, 19.61) | 20.86 (16.16, 25.55) | 12.96 (9.10, 16.62) | 16.97 (5.22, 28.71) | 16.96 (13.20, 20.73) | 17.87 (12.75, 23.00) | 10.93 (2.24, 19.62) |
| <14,999 | 216 | 28.28 (24.81, 31.75) | 34.07 (28.66, 39.48) | 20.73 (26.09, 25.36) | 43.27 (28.42, 58.12) | 29.86 (25.25, 34.48) | 27.43 (21.48, 33.39) | 19.20 (8.84, 29.56) |
| 15-29,999 | 160 | 21.37 (18.20, 24.53) | 24.99 (19.97, 30.00) | 20.19 (15.61, 24.80) | 12.73 (4.02, 21.44) | 21.07 (16.96, 25.19) | 18.42 (13.32, 23.49) | 37.62 (23.58, 51.66) |
| 30-49,999 | 123 | 16.25<br>(13.39, 19.11) | 11.47 (7.71,<br>15.23) | 20.72 (16.20,<br>25.23) | 14.40 (3.14,<br>25.67) | 17.63 (13.70,<br>21.56) | 15.22 (10.51,<br>19.93) | 11.22 (3.12,<br>19.33) |
| 50-79,999 | 93 | 10.72<br>(8.50, 12.93) | 4.84 (2.36,<br>7.32) | 16.14 (12.37,<br>19.92) | 9.98 (1.85,<br>18.12) | 7.60 (5.19,<br>10.00) | 15.67 (11.29,<br>20.06) | 13.17 (2.46,<br>23.88) |
| 80,000+ | 53 | 6.65<br>(4.78, 8.53) | 3.77 (1.69,<br>5.86) | 9.36 (6.18, 12.54) | 2.65 (0, 6.44) | 6.87 (4.45, 9.30) | 5.39 (2.44, 8.33) | 7.85 (.27, 15.43) |
| Education |  |  |  |  |  |  |  |  |
| < High school | 25 | 5.57 (3.39, 7.75) | 7.42 (3.65,<br>11.20) | 3.48 (1.01, 5.95) | 3.88 (0, 11.27) | 5.52 (2.60, 8.44) | 4.46 (1.93, 7.73) | 10.78 (.83,<br>20.73) |
| High school diploma | 64 | 11.16 (8.49, 13.83) | 12.74 (8.4,<br>17.08) | 11.00 (7.15,<br>14.85) | 1.68 (0, 4.97) | 12.50 (8.83,<br>16.17) | 9.85 (5.38, 14.31) | 7.58 (.26,<br>14.895) |
| Some college or university | 218 | 27.02 (24.26, 30.97) | 28.81 (23.77,<br>33.85) | 24.40 (19.61,<br>29.19) | 41.70 (27.22,<br>56.17) | 27.57 (23.20,<br>31.95) | 29.09 (23.24,<br>34.94) | 20.35 (9.08,<br>31.62) |
| College or university degree | 343 | 42.14 (38.41, 45.87) | 41.99 (36.40,<br>47.58) | 43.61 (38.12,<br>49.11) | 39.93 (25.63,<br>54.22) | 40.44 (35.58,<br>45.30) | 43.00 (36.61,<br>49.39) | 50.71 (36.81,<br>64.61) |
| Grad/professional degree | 130 | 13.51 (11.13, 15.89) | 9.04 (6.03,<br>12.06) | 17.51 (13.69,<br>21.33) | 12.82 (2.37,<br>23.27) | 13.96 (10.86,<br>17.06) | 13.61 (9.35,<br>17.86) | 10.58 (2.72,<br>18.44) |

a. Calculated using Statistics Canada 2019 low-income measure, which adjusts household income by number of people supported by income.

### Mental health

#### Depression

Pre-pandemic and pandemic depression symptoms were assessed using the 10-item Center for Epidemiologic Studies Depression Scale (CES-D-10) [17]. Items are rated in frequency on a scale of 1 (*rarely or none of the time, less than 1 day*) to 4 (*most or all of the time, 5–7 days*). Final scores are calculated by reverse-scoring as needed and adding item ratings (range: 0-30). A final score of 10 or higher is generally used as a cut-off for significant depression symptoms [17]. Internal consistency was high at pre-pandemic baseline (Cronbach’s alpha, α = .86) and at pandemic follow-up (α = .86).

#### Anxiety

Baseline and pandemic anxiety were assessed using the 5-item Overall Anxiety Severity and Impairment Scale (OASIS) [18]. Items are ranked in severity on a 5-point scale (1= *Not at all*, 5=*Extreme*) and summed (range: 0-20). A total score of 8 or higher is generally used as a cut-off for significant anxiety symptoms [18]. Internal consistency was high in the present sample at pre-pandemic baseline (α = .89) and pandemic follow-up (α = .88).

#### Social Support and Living Situation

We assessed social support using the 8-item Medical Outcomes Study Social Support Scale (MOS-SS) [19]. Items are rated in frequency on a 5-point scale (*Never* to *Constantly*). The MOS-SS has two subscales, Tangible and Emotional support, which can be considered separately or as a whole. In our study, we averaged all 8 items for an overall score, with higher scores indicating more social support. The MOS-SS had high internal consistency in this sample at pre-pandemic baseline (α = .91). Participants were also asked if conditions related to COVID had caused them to live with a household member who is unsupportive of their gender (*1 = Yes, 0 = No*).

#### Change in Finances Due to COVID-19

To create a composite item for financial change during COVID, we combined two items from the pandemic follow-up survey. Both items were adopted from Statistics Canada surveys [20]. The first of these items asked participants to rank the impact COVID-19 had on their ability to meet financial obligations or essential needs (e.g., rent, mortgage payments, utilities, groceries). The second item asked participants to rank the impact that COVID-19 had on their personal income from employment. Both of these items were ranked on an 8-point scale (*major negative impact, moderate negative impact, minor negative impact, no impact, minor positive impact, moderate positive impact, major positive impact, too soon to tell*). When combining these items, *too soon to tell* was recoded as a missing value. We then recoded these two items into one composite item using the following guidelines: if participants reported a negative major impact on either item (regardless of the response on the other item), they were coded with 0 to represent *negative change*; if participants reported minor positive or negative impact or no change on both items, they were coded as *no change* (1). If participants reported moderate or major positive impact on both items, they were coded as *positive change* (2). If participants reported neutral or minor change on one item and moderate or major positive impact on another, they were coded as *positive change* (2). If participants reported minor or no change on one item and did not respond to the second item, they received a missing score.

#### Access to Trans and Non-binary Peer or Friend Gatherings

In the pandemic follow-up survey, we asked participants to rank how their access to TNB peer or friend gatherings (online or in person) had changed since March 2020. This question was rated on a 3-point scale (*decreased, stayed the same, increased*).

## Statistical Analysis

### Missing data

The percentage of missing values in relevant variables ranged from 0 to 6% of cases and relevant data was complete for 84.6% of participants. The pattern of missing data was determined to be non-monotone, suggesting that missing data was not due to issues related to participant attrition during survey completion. For multivariable regression analyses, we conducted multiple imputation using fully conditional specification in SAS 9.4, producing 20 imputations. Outcome variables (pandemic depression and anxiety) were not imputed. Sensitivity analyses revealed similar outcomes when analyses were run with the Missing *Not At Random* command. Similar results were obtained when analyses were restricted to complete case data. Descriptive data (Table 1) were obtained prior to imputation.

### Survey weights and descriptive analyses

We weighted all analyses using Stata ipfweight, which performs stepwise adjustment to achieve known population margins. Our weights represented the full sample, including the short version-only respondents to the 2019 survey, to ensure that results were representative of the full study sample. Subsequent analyses were performed using survey procedures in SAS 9.4. We estimated weighted frequencies and used t-tests to assess changes in depression and anxiety from pre-pandemic baseline to pandemic follow-up, both using non-imputed data.

### Multivariable linear regression

We fit four multivariable regression models to examine the relationship between financial change and mental health (depression and anxiety symptoms) and then change in access to TNB peer or friend gatherings and mental health. In SAS 9.4, PROC MIANALYZE and PROC SURVEYREG were used to combine the 20 imputed datasets and weight regression analyses after imputation. R^2^ and adjusted R^2^ were obtained by averaging output from the 20 imputations. Prior to analysis, we checked regression diagnostics and determined that 2019 income-to-needs ratio and pre-pandemic social support were skewed. Therefore, in a sensitivity analysis, we conducted regression analyses with these log-transformed variables. In all cases, these transformed analyses showed the same patterns of results. We thus present the analyses without transformation.

### Confounders

Confounders were selected using directed acyclic graphs in DAGitty.net [21, 22]. Previous research and community knowledge were used to select variables that impact mental health, as well as factors that influence those variables. In models containing financial change as the exposure, confounders (measured at pre-pandemic baseline) were age, gender, immigration history (yes/no), employment, education, income-to-needs ratio, indigeneity (yes/no), province, and racialization (yes/no). In models with change in trans and non-binary gathering access as the exposure, confounders were sex assigned at birth, province at pre-pandemic baseline, racialization, gender, immigration history, pre-pandemic social support, and living with an unsupportive household member during the pandemic (yes/no).

## Results

### Sample characteristics and descriptive statistics

Demographic characteristics of the sample are presented in Table 1, stratified by changes in finances and access to TNB peer or friend gatherings. The mean age of the sample was 33.0 years (95% CI 32.12, 33.84, range: 18-75). Over half of our sample reported that COVID had a major (9.17%, 95% CI 6.88, 11.46), moderate (20.27%, 95% CI 17.22, 23.33), or minor (24.05%, 95% CI 20.76, 27.35) negative impact on their ability to meet financial obligations or essential needs, and 30.08% (95% CI: 26.57, 33.58) reported no change. Some participants reported a major (2.51%, 95% CI 1.27, 3.74), moderate (6.00%, 95% CI 4.06, 7.93), or minor (7.92%, 95% CI 5.75, 10.09) positive change in ability to meet financial obligations or essential needs. Approximately half of the sample reported that COVID caused major (17.61%, 95% CI 14.68, 20.54), moderate (16.36%, 95% CI 13.54, 19.18), or minor (15.95%, 95% CI 13.10, 18.79) negative impact on their personal income, and 37.50% (95% CI 33.77, 41.24) reported no change. The remaining participants reported a major (1.80%, 95% CI .61, 2.99), moderate (3.41%, 95% CI 2.02, 4.80), or minor (7.38%, 95% CI 5.32, 9.54) positive impact on income. 58.3% (95% CI 54.52, 62.03) of participants reported that they experienced loss in access to trans and non-binary peer or friend gatherings since March 12, 2020, while 33.58% (95% CI 30.00, 37.17) reported no change and 8.14% (95% CI 5.98, 10.30) reported an increase. During the pandemic, most participants (72.1%) were above the CESD cut-off for depression, and 76.6% were above the OASIS cut-off for anxiety symptoms. Mean depression scores increased significantly from pre-pandemic baseline (*M* = 14.47, 95% CI 13.93, 15.01) to pandemic follow-up (*M* = 16.50, 95% CI 15.98, 17.01), *t*(730) = 9.04, *p* <.0001. Mean anxiety scores also increased significantly from pre-pandemic baseline (*M* = 10.13, 95% CI 9.80, 10.46) to pandemic follow-up (*M* = 10.35, 95% CI 10.04, 10.66), *t*(730)=2.29, *p* = .022.

### Multivariable regression analysis

#### Association between change in finances and pandemic depression (Table 2)

There was a significant interaction between changes in finances and pre-pandemic depression (*p* = .0261). Here, we found that, for every unit increase in pre-pandemic depression, participants with no change in finances experienced greater increase in pandemic depression than participants reporting loss in finances. In other words, individuals with low pre-pandemic depression and financial stability (no loss or gain) report pandemic depression that was comparable to those experiencing financial increase; however, participants with high pre-pandemic depression and no financial change reported pandemic depression that was comparable to individuals reporting financial loss (Fig. 1) This first model explained 38% of the variance in depression.

**Fig. 1.**
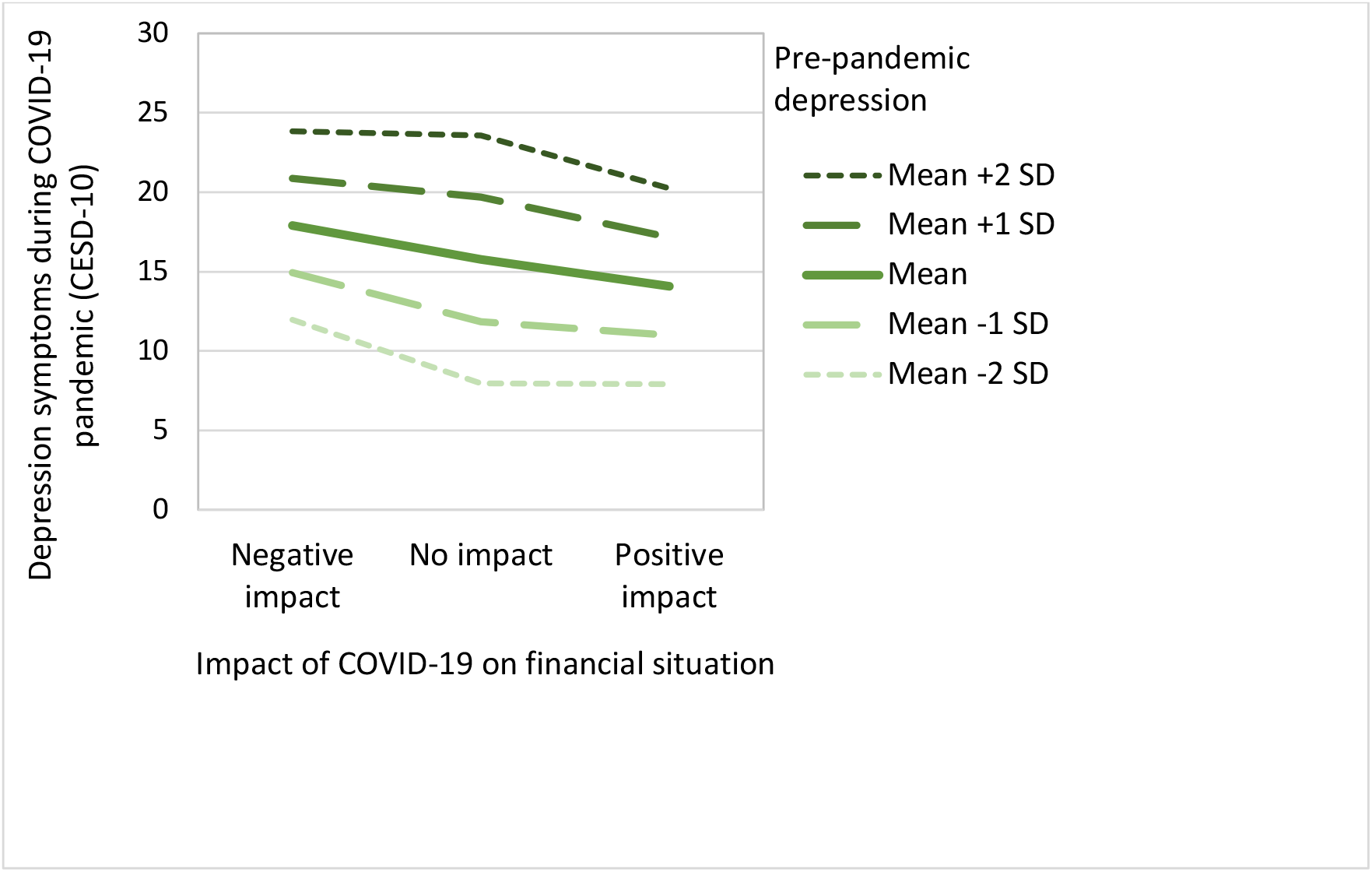
Pandemic depression symptoms by COVID-19’s impact on finances and pre-pandemic depression symptoms

**Table 2.** Linear regression predicting mental health in COVID with financial change during the pandemic (*n* = 780)

|  | **Outcome: Depression symptoms during pandemic (CESD)** |  |  | **Outcome: Anxiety symptoms during pandemic (OASIS)** |  |  |
| --- | --- | --- | --- | --- | --- | --- |
|  | Coefficient | 95% CI | *p* value | Coefficient | 95% CI | *p* value |
| Intercept | 14.10 | 10.58, 17.61 | <.0001 | 5.52 | 3.30, 7.74 | <.0001 |
| Financial Change |  |  |  |  |  |  |
| Negative | Reference |  |  | Reference |  |  |
| No Change | -4.11 | -7.95, -.051 | <.0001 | 1.15 | -2.68, .38 | .140 |
| Positive | -4.07 | -6.12, -.21 | .0425 | 1.02 | -1.71, 3.74 | .465 |
| Baseline CESD | .43 | .34, .53 | <.0001 | -- | -- | -- |
| Baseline OASIS | -- | -- | -- | .59 | .49, .69 | <.0001 |
| Baseline CESD x Financial Change |  |  |  | -- | -- | -- |
| CESD x Negative | Reference |  |  |  |  |  |
| CESD x No Change | .14 | .02, .26 | .0253 |  |  |  |
| CESD x Positive | .02 | -.24, .27 | .818 |  |  |  |
| Baseline OASIS x Financial Change | -- | -- | -- |  |  |  |
| OASIS x Negative |  |  |  | Reference |  |  |
| OASIS x No Change |  |  |  | .05 | -.08, .18 | .426 |
| OASIS x Positive |  |  |  | -.21 | -.50, .08 | .154 |
| $R^2$ | .40 | | | .44 | | |
| Adjusted $R^2$ | .38 | | | .41 | | |

All models control for age, gender, immigration status (yes/no), racialization (yes/no), Indigeneity (yes/no), 2019 income-to-needs ratio, province, baseline education, baseline employment.
CESD, Center for Epidemiological Studies Depression; OASIS, Overall Anxiety Severity and Impairment Scale.

#### Association between change in finances and pandemic anxiety

In examining pandemic anxiety, we found no interaction with pre-pandemic anxiety. Change in finances did not significantly contribute to the model, but pre-pandemic anxiety did (*p* < .0001). This model accounted for 41% of variance in pandemic anxiety.

#### Association between change in trans and non-binary gathering access and pandemic depression (Table 3)

There was a significant interaction between change in access to trans and non-binary peer or friend gatherings and pre-pandemic depression (*p* = .0005) such that for every unit increase in pre-pandemic depression, participants reporting an increase in trans and non-binary gathering access experienced significantly less pandemic depression than individuals reporting decrease in gathering access or no change in gathering access. Participants who reported low pre-pandemic depression symptoms and an increase in access to TNB peer or friend gatherings reported higher pandemic depression than their counterparts who reported decrease or no change in gathering access. Conversely, individuals with high pre-pandemic depression and increased TNB gathering access reported less pandemic depression than those who reported decreased gathering access. Participants with high pre-pandemic depression who reported increase in TNB gathering access appeared to show less pandemic depression than people who had high pre-pandemic depression and reported either loss in access or no change in access (Fig. 2). This model predicted 28% of the variance in pandemic depression symptoms.

**Fig. 2.**
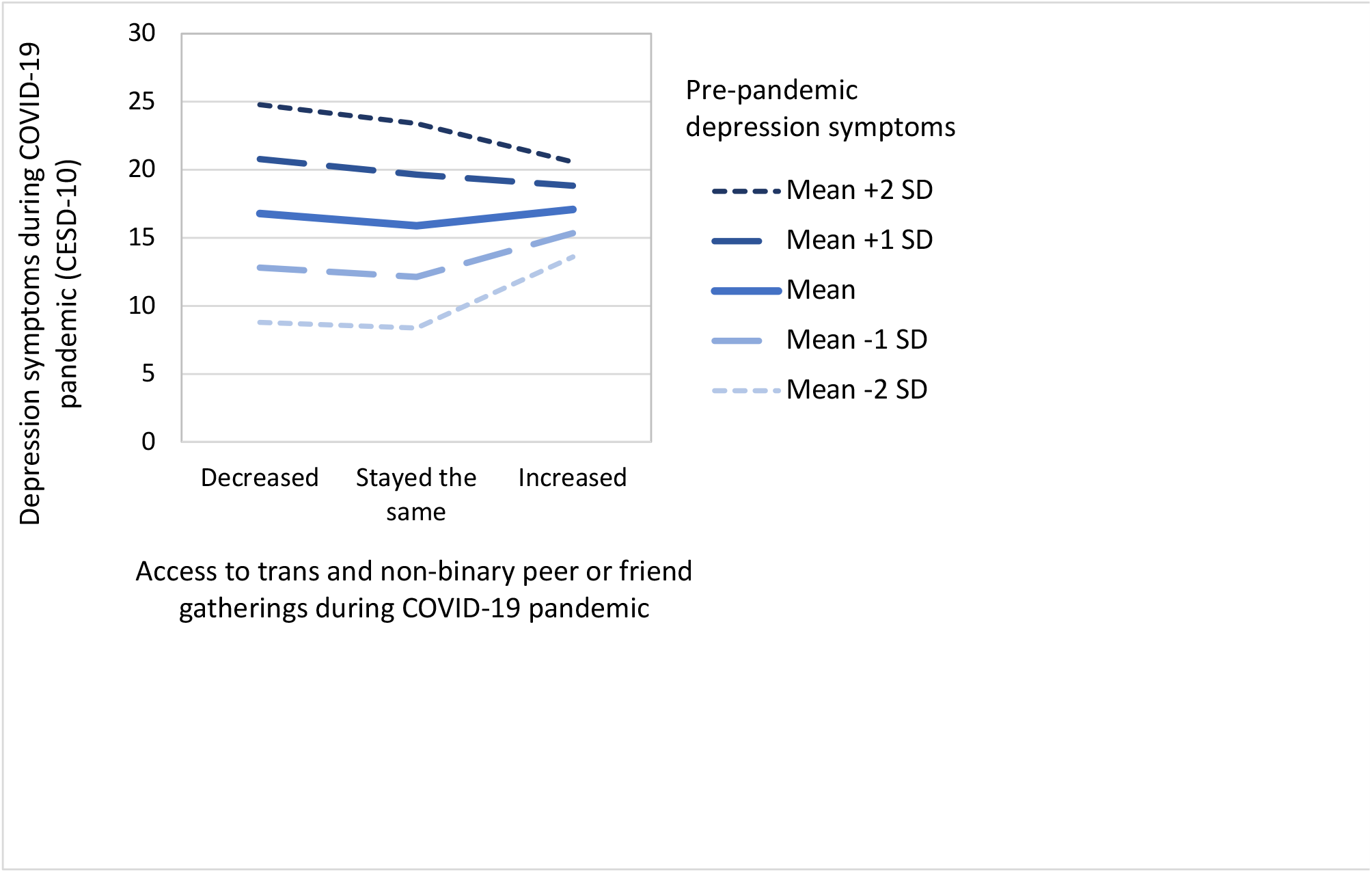
Pandemic depression symptoms by change in trans and non-binary gathering access during COVID-19 and pre-pandemic depression symptoms

**Table 3.** Linear Regression Predicting Mental Health in COVID with Change in Trans and Non-binary Gathering Access (*n* = 780)

|  | **Outcome:** Depression symptoms during pandemic |  |  | **Outcome:** Anxiety symptoms during pandemic |  |  |
| --- | --- | --- | --- | --- | --- | --- |
|  | Coefficient | 95% CI | *p* value | Coefficient | 95% CI | *p* value |
| Intercept | 9.41 | 6.32, 12.51 | <.0001 | 14.93 | 11.68, 18.18 | <.0001 |
| Change in Gathering Access |  |  |  |  |  |  |
| Decrease | Reference |  |  | Reference |  |  |
| No Change | -.41 | -2.44, 1.63 | .696 | -1.00 | -3.84, 1.84 | .489 |
| Increase | 5.03 | 2.05, 8.01 | .0009 | 1.74 | -2.73, 6.20 | .446 |
| Baseline CESD | .58 | .50, .67 | <.0001 | --- | --- | --- |
| Baseline OASIS | -- |  |  | .68 | .51, .85 | <.0001 |
| Baseline CESD x Change in Gathering Access |  |  |  |  |  |  |
| CESD x Decrease | Reference |  |  | Reference |  |  |
| CESD x No Change | -.034 | -.16, .09 | .597 |  |  |  |
| CESD x Increase | -.33 | -.51, -.15 | .0004 |  |  |  |
| Baseline OASIS x Change in Gathering Access | --- | --- | ---- |  |  |  |
| OASIS x Decrease |  |  |  | Reference |  |  |
| OASIS x No Change |  |  |  | .04 | -.22, .31 | .75 |
| OASIS x Increase |  |  |  | -.07 | -.48, .35 | .77 |
| $R^2$ | | .30 | | | .52 | |
| Adjusted $R^2$ | | .28 | | | .50 | |

All models control for gender, immigration status (yes/no), racialization (yes/no), province, baseline social support, sex assigned at birth (male/female), report of living with unsupportive household member (yes/no).
CESD, Center for Epidemiological Studies Depression; OASIS, Overall Anxiety Severity and Impairment Scale.

#### Association between change in trans and non-binary peer or friend gathering access and pandemic anxiety

We found no interaction between pre-pandemic anxiety symptoms and change in trans and non-binary gathering access. Change in trans and non-binary peer or friend gathering access did not significantly contribute to the model with pandemic anxiety as an outcome, but pre-pandemic anxiety did (p < .0001). This final model predicted 50% of variance in pandemic anxiety.

## Discussion

We examined associations between changes in finances and TNB peer or friend gathering access and mental health during the COVID-19 pandemic. Approximately half of our sample reported a negative change in finances due to the pandemic. The majority reported a loss in access to TNB peer or friend gatherings (online and in person). We found that both depression symptoms and anxiety severity/impairment increased significantly in our sample from pre-pandemic baseline to pandemic follow-up, and that most participants were above cut-offs for both depression and anxiety during the pandemic. When observing pandemic depression symptoms, we found that the impact of changes in finances and TNB gathering access varied depending on pre-pandemic depression, but that change in finances and TNB gathering access did not predict pandemic anxiety in our sample.

Our findings on the impact of financial change on pandemic depression substantiate previous research showing that income loss during COVID-19 is linked with reduced mental health among TNB people [23] and that pre-pandemic mental health predicts pandemic mental health [24, 25, 26]. Our results, however, advance the research on these associations and reveal an interaction between pre-pandemic depression and financial loss. For individuals low in depression symptoms prior to the pandemic, financial stability was protective; meanwhile, for individuals who entered the pandemic with high depression symptoms, financial stability was not protective, and depression symptoms increased at a greater rate during the pandemic. Past research has also noted complex associations between mental health and finances. For example, research on employment security and mental health of low-income, urban women showed that employment security was only protective of mental health if the women had access to childcare [27]. In other words, financial stability does not directly improve mental health, and the relationship between financial status and mental health is complicated by other factors (e.g., tangible support).

We also found that the relationship between access to peer gatherings and pandemic depression varied based on pre-pandemic depression. Participants with low pre-pandemic depression and increase in TNB gathering access reported higher pandemic depression than those with low pre-pandemic depression and a decrease or no change in community access. Individuals reporting high pre-pandemic depression and increased access to TNB peer or friend gatherings reported reduced pandemic depression compared to individuals high in pre-pandemic depression who experienced loss in community access. It is possible that participants with low pre-pandemic depression pursued more community connection/access during the pandemic because they were experiencing increased depression symptoms. Thus, for those with low pre-pandemic depression, increased depression may have led to increased gathering access. Meanwhile, individuals who entered the pandemic with high depression and reported an increase in gathering access experienced the reduced depression that would be expected to result from an increase in community engagement. Supportive of our findings, research with a nationally representative sample in the United States has shown that building social trust with surrounding community was protective for people with major depression at pre-pandemic baseline, but this effect was not found for those without depression at pre-pandemic baseline [28]. Results from the same study showed that, while trust was protective for people with pre-pandemic depression, community participation was not – suggesting that merely having access to community may not be protective and highlighting the importance of separating social capital into different dimensions when examining the relationship with mental health [28]. When examining the relationship between social support and resilience in the general population, research has suggested that individuals with low resilience need high levels of support from family, friends, and community for mental health to be protected from stress [29].

We found that neither change in finances nor in TNB gathering access were significantly associated with levels of pandemic anxiety. Consistent with previous research, pre-pandemic anxiety was the primary predictor of follow-up anxiety (e.g., [30]) and, specifically, pandemic anxiety [31]. In research conducted prior to COVID-19 that predicted follow-up mental health from housing insecurity and baseline mental health, results revealed that the predictive power of housing insecurity was reduced when controlling for baseline mental health [32]. This has also been found more specifically in predictions of anxiety during COVID [33].

Our results show that anxiety persists, regardless of change in finances or TNB gathering access, whereas pandemic depression may be assuaged or enhanced. This is in line with recent COVID-related research which showed that anxiety increased during the pandemic, but depression did not [34]. Prior to COVID-19, other research has shown that anxiety tends to persist more than depression or other forms of mental distress. For example, when examining the relationship between social support and changes in mental health among heart failure outpatients, research has shown that increased social support contributed to changes in depression but did not affect anxiety [35]. In a study of anxiety and distress in patients with multiple sclerosis, Janssens et al. [36] found that distress decreased over time, but that high levels of anxiety did not change. These authors attributed this to uncertainty and concerns about future consequences of patient health. Uncertainty is likely a major contributor to anxiety or distress during COVID and has been linked to increased pandemic eating disorder pathology [37]. Uncertainty may thus be one reason that anxiety persisted in our sample, regardless of change in finances or access to TNB peer or friend gatherings.

Although our study had many strengths, including a large and diverse national sample, as well as pre-pandemic assessments of mental health, there were limitations. Our measure of access to TNB peer or friend gatherings did not specify the nature of this access. Findings on the relationship between gathering access and mental health in the literature vary based on context. In some cases, among marginalized groups, high bonding with similar others is associated with worsened mental health, whereas connecting to individuals who are socio-demographically different is associated with less mental distress [38]. If the communities that participants were accessing are related to activism and civic engagement, there are mixed findings on the benefits of that type of engagement on mental health (e.g., [39]). Access to gatherings may also have been in the form of social media “lurking,” which may have a negative relationship with perceived social support and be related to worsened mental health [40]. This limits the interpretability of our findings. Qualitative investigation (e.g., [41]) may provide a deeper understanding of different dimensions of peer or friend gathering access and the impact on mental health. We additionally were unable to include participants who did not complete the COVID-19 follow-up survey and it is possible that their unavailability was related to distress or financial strain.

## Conclusion

The present study used prospective data to assess mental health during the COVID-19 pandemic and the impact that changes in finances or trans and non-binary peer or friend gathering access has on depression and anxiety. Overall, we found that the relationship between changes in finances and access to TNB gatherings and pandemic depression varied, depending on pre-pandemic depression – but that these moderating effects were not found when examining pandemic anxiety. Thus, although depression varied, anxiety persisted and increased, regardless of financial or gathering access change. This may have been due to the general uncertainty experienced during COVID; even with, for example, stability in finances or increase in access to TNB gatherings, there are many areas of uncertainty brought about by the COVID-19 pandemic. Our findings suggest that minor or temporary financial assistance is likely an oversimplified solution to increased mental health concerns during the pandemic and that TNB individuals may benefit from more meaningful resources and aids that address different dimensions of need. Prior to and during COVID, the sample average in depression and anxiety metrics surpassed the cut-offs indicating potential mental health disorders – and pre-pandemic mental health was found to predict pandemic mental health very strongly. This highlights the insufficiency of temporary financial aid during the pandemic and indicates the importance of developing an established, multifaceted approach to mental health programmes and services to address structural barriers and needs, including financial support and meaningful community engagement. Such programs may also help to reduce COVID-related and general uncertainty, and thus alleviate anxiety.

## Data Availability

The 2019 Trans PULSE Canada survey data cannot be deposited in a data repository due to the Research Ethics Board approval and conditions in the letter of information and consent for participants that specified data would only be seen by members of the large national team. These conditions were necessary to build this community based research project in the context of stigmatization and distrust of researchers. The more limited 2020 COVID data subset are available through an interactive data dashboard at transpulsecanada.ca/covid/. A larger version of the 2020 data set will be deposited in a repository prior to publication.

## Statements & Declarations

### Availability of data and material

The 2019 Trans PULSE Canada survey data (baseline data source) cannot be deposited in a data repository due to the Research Ethics Board approval and conditions in the letter of information and consent for participants that specified data would only be seen by members of the large national team’s Data Analysis Working Group. These conditions were necessary to build this community-based research project in the context of stigmatization and distrust of researchers. For the more limited 2020 COVID-specific data subset (follow-up/pandemic data source), data are available through an interactive data dashboard at https://transpulsecanada.ca/covid/. A larger version of the 2020 data set is being cleaned and deidentified and will be deposited in a repository prior to publication.

### Code availability

N/A

### Ethics approval

All procedures performed in studies involving human participants were in accordance with the ethical standards of the institutional and/or national research committee and with the 1964 Helsinki Declaration and its later amendments or comparable ethical standards. Trans PULSE Canada was approved by the research ethics boards of Western University, Unity Health Toronto, Wilfrid Laurier University and the University of Victoria; the TPC COVID Cohort follow-up was approved at Western University, Wilfrid Laurier University, University of Victoria, Universal Health Network, University of British Columbia, and Drexel University.

### Consent

Informed consent was obtained from all individual participants included in the study. Participants consented to publication of results from this research.

### Authors’ contribution statements

MG: Formal data analysis, substantial interpretation of results, writing – original draft, writing – review and editing

AS: Original study design, survey design, data collection, writing – review and editing

CC: Survey design, data collection, writing – review and editing

HS: Drafted introduction (writing – original draft)

NA: Survey design, writing – review and editing

GB: Original study design, survey design, data collection, data analysis, figures, writing – review and editing

## Acknowledgments

The Trans PULSE Canada Study Team would like to acknowledge and thank the trans and non-binary people who have generously shared their time and experience with us. The Trans PULSE Canada Study was funded by the Canadian Institutes of Health Research (Funding Reference Number PJT-159690).

